# A Mixed-Methods Investigation of Coping, Adaptation and Health-Related Quality of Life in Individuals Experiencing Endometriosis

**DOI:** 10.1101/2024.02.19.24303022

**Authors:** Chloe Moore, Nicola Cogan, Lynn Williams

## Abstract

**Objectives:** Endometriosis is associated with reduced health-related quality of life (HRQoL). Coping has been linked to HRQoL in this population but longitudinal research to establish the impact of coping on HRQoL over time is lacking. Additionally, limited research has examined coping and adaptation to endometriosis from an in-depth qualitative or mixed-methods perspective. Therefore, adopting a mixed-methods approach, the current research aimed to investigate how individuals experiencing endometriosis coped and adapted to their condition, and the extent to which coping predicted HRQoL over time.

**Design:** A sequential, mixed-methods design incorporating a two-wave longitudinal survey and semi-structured interviews.

**Methods:** 408 UK-based participants diagnosed with endometriosis completed the baseline survey, measuring demographics, clinical factors, and coping with 283 completing the follow-up survey assessing HRQoL a year later. Data was analysed through hierarchical regression analysis. Meanwhile, 30 semi-structured interviews were conducted and analysed using reflexive thematic analysis. Findings were integrated by considering points of convergence, divergence, and complementarity between the datasets.

**Results:** Analysis of the quantitative data indicated an enduring impact of coping on HRQoL. Avoidant strategies and endometriosis-related information-seeking predicted reduced HRQoL, whilst trust in medical care exerted a protective function. Three themes were constructed from the qualitative data: disconnection from the body; balancing boundaries and self-care; and empowered adaptation. Integration of the datasets emphasised the significant impact of coping on HRQoL and wellbeing, revealing avoidance and positive adaptation as prominently employed coping strategies.

**Conclusions:** The results position coping as an important therapeutic target in endometriosis care, particularly through fostering empowerment and adaptation to support HRQoL.

## Introduction

Endometriosis is a chronic, incurable condition in which tissue similar to the lining of the womb, the endometrium, is found outside the uterus (Zondervan et al., 2020). Symptoms vary between individuals, however, common complaints include chronic pelvic pain, dyspareunia, and heavy menstrual bleeding (World Health Organization, 2023). One in ten women and individuals assigned female at birth experience endometriosis (World Health Organization, 2023).

Endometriosis is associated with reduced quality of life (QoL) and a heightened risk of depression and anxiety compared to the general population (Wang et al., 2021). Prevalence of depression and anxiety in this population has been placed as high as 86.5% and 87.5% respectively (Sepulcri & do Amaral, 2015). The severe and often debilitating symptoms experienced by many with the condition are thought to underlie adverse QoL outcomes.

Specifically, pain has been linked to reduced QoL in the context of endometriosis (Rees et al., 2022). As pain increases, so too does the risk of depression, anxiety, and adverse QoL outcomes (Facchin et al., 2015). However, the progressive and incurable nature of endometriosis means that, for many, pain treatment is limited in its effectiveness (Carlyle et al., 2020), with 40-50% of diagnosed individuals experiencing symptom recurrence within 5 years following surgery to remove endometriosis lesions (Horne & Saunders, 2019). Therefore, the contribution of alternate factors to QoL have been considered, with the intention of mitigating the detrimental impact of endometriosis symptomology on QoL and wellbeing outcomes.

Negative body image (Van Niekerk et al., 2022), diagnostic delays (Gallagher et al., 2018), and sub-fertility (Vitale et al., 2017) have all been linked to endometriosis-related QoL, suggesting that QoL in this population is determined by a complex interplay between several physiological, social and psychological factors. Such factors may influence QoL directly, or indirectly through driving coping responses.

Coping is thought to play a pivotal role in shaping endometriosis-related QoL and wellbeing outcomes (Zarbo et al., 2018). Specifically, avoidant coping strategies are linked to worsened mental health and wellbeing, whilst adaptive coping strategies are associated with more positive wellbeing outcomes (Eriksen et al., 2008; González-Echevarría et al., 2019). In a review of the existing literature, Zarbo et al. (2018) described several coping strategies adopted by individuals experiencing endometriosis, including emotion suppression, pain catastrophising, seeking social support, and self-management. Emotional and avoidant coping styles predicted adverse mental health outcomes, whilst adaptive, rational coping styles were related to more positive mental health effects.

Qualitative research has also considered the specific coping styles employed by individuals experiencing endometriosis. For example, Grogan et al. (2018) observed coping strategies such as hiding endometriosis symptoms, rejecting painkillers, and self-pacing through avoiding social events and prioritising work commitments to be prominent within this population. The impact of these coping styles on wellbeing was complex. For instance, concealing symptoms allowed a sense of normality and often had a protective effect on the self-concept, however coping in this way also led to feelings of guilt and isolation, negatively impacting participants’ relationships. The authors stress the necessity for tailored support unique to each individual, which may in turn promote adaptation to enable individuals with endometriosis to live a fulfilling life.

Outwith endometriosis, researchers have identified a link between disease acceptance and positive HRQoL outcomes in several chronic illnesses, such as chronic obstructive pulmonary disease (Jankowska-Polańska et al., 2016) and chronic heart failure (Obieglo et al., 2016). Disease acceptance is associated with improved adaptation and reduced emotional distress (Wysocki et al., 2023), highlighting the potential benefits of positive adaptation in chronic conditions such as endometriosis.

However, there is a need for increased understanding of way in which coping and adaptation impact the lives of individuals with endometriosis before recommendations for psychological interventions can be made. Although existing research strongly indicates that coping impacts QoL and wellbeing in this population, notably missing from the literature is longitudinal work to ascertain the long-term predictive validity of coping strategies on endometriosis-related QoL outcomes. Additionally, there is limited exploration of the specific coping strategies employed by individuals with endometriosis from an experiential or mixed-methods perspective. Therefore, adopting a mixed-methods approach, the current research aims to combine qualitative and quantitative insights relating to the way in which individuals with endometriosis cope with and adapt to their condition. Specifically, the current study firstly aims to establish whether a range of health-directed coping strategies predict health-related QoL (HRQoL) in endometriosis over the course of one year. Secondly, a qualitative component aims to gain an in-depth understanding of the coping and adaptation processes adopted by individuals experiencing the condition. Findings from each element of the current study will be combined, by considering points of overlap and divergence amongst the datasets to produce an in-depth account of adaptation and coping in this population.

## Methods

### Research Design

A mixed methods sequential design was adopted, incorporating in-depth, semi-structured interviews and a two-wave longitudinal survey. The two components of the current research were analysed separately before the findings were integrated. Ethical approval was obtained from the University’s ethics committee (ethics code: UEC21/40). Data collection occurred between April 2021 and June 2023.

### Longitudinal Survey

#### Participants

Four hundred and eight participants were recruited for the baseline (T1) survey through convenience sampling. Participants were largely recruited from endometriosis support organisations and social media. Individuals over the age of 18, residing in the UK or Ireland, and who self-reported a medically confirmed diagnosis of endometriosis were eligible to participate. Twelve months after the baseline survey, 283 participants completed the follow-up (T2) survey.

T-tests and chi-square analyses revealed no significant differences between those who completed the follow-up survey and those who did not in demographic background or health status.

At T1, participants were aged between 19 and 56 (M = 33.92; SD = 7.99). Details regarding gender were not recorded, although it is important to note that not everyone who experiences endometriosis identifies as female. Further demographic and clinical details can be found in table 1.

**Table 1.** Participant demographics and clinical information at T1.

|  | N | % |
| --- | --- | --- |
| **Ethnicity** ($n = 408$) | | |
| White English | 159 | 39% |
| White Scottish | 122 | 29.9% |
| Another White Background | 34 | 8.3% |
| White Welsh | 34 | 8.3% |
| White Irish | 21 | 5.1% |
| White Northern Irish | 15 | 3.7% |
| Asian and White | 5 | 1.2% |
| Indian | 4 | 1.0% |
| African | 4 | 1.0% |
| Another Mixed Background | 4 | 1.0% |
| Pakistani | 2 | 0.5% |
| Other | 1 | 0.2% |
| Another Asian Background | 1 | 0.2% |
| Another Ethnic Background | 1 | 0.2% |
| Caribbean | 1 | 0.2% |
| **Educational attainment** ($n = 407$) | | |
| Attended university | 251 | 61.7% |
| Did not attend university | 156 | 38.3% |
| **Employment status** ($n = 406$) | | |
| Employed full-time / self-employed | 242 | 59.6% |
| Employed part-time / casual contract | 70 | 17.2% |
| Student | 36 | 8.9% |
| Unemployed | 30 | 7.4% |
| Disabled or unable to work | 28 | 6.9% |
| **Country of residence** ($n = 408$) | | |
| England | 187 | 45.8% |
| Scotland | 147 | 36% |
| Wales | 43 | 10.5% |
| Northern Ireland | 16 | 3.9% |
| Ireland | 15 | 3.7% |
| **Co-morbid condition** (*n* = 401) |  |  |
| Yes | 240 | 59.9% |
| No | 161 | 40.1% |
| **Surgery** (*n* = 407) |  |  |
| Had surgery | 343 | 84.3% |
| Not had surgery | 64 | 15.7% |

Participants had experienced endometriosis for approximately 15.5 years (SD = 8.44) and had been diagnosed for around 5.1 years (SD = 5.61), indicating a mean diagnostic delay of 10.4 years.

#### Materials

The current study was conducted as part of a larger project examining illness perceptions in endometriosis (Moore et al., 2023). Only the measures relevant to the aims of the current paper will be described.

#### Demographics

At T1, participants provided demographic and clinical data, including ethnicity, educational attainment, employment status, household income, country of residence, whether they experienced co-morbid conditions, their history of endometriosis surgery, and the duration of their endometriosis symptoms and diagnosis.

#### Coping

Also at T1, participants completed the 45-item Essen Coping Questionnaire (Franke & Jangla, 2016), which measures 9 coping styles specific to experiencing ill-health/disease: 1) acting, problem-orientated coping; 2) distance and self-promotion; 3) information seeking and exchange of experiences; 4) trivialisation, wishful-thinking and defence; 5) depressive processing; 6) willingness to accept help; 7) active search for social integration; 8) trust in medical care; 9) finding inner stability. Each item is ranked on a 5-point Likert scale (0: ‘Not at all’; 5: ‘Extremely’). Scores for each category of coping were derived by summing up the responses to the corresponding questions and dividing them by the number of questions to gain an average for each coping style. The reliability of each subscale is presented in table 2. Acting, problem-orientated coping, information seeking and exchange of experiences, depressive processing, and trust in medical care all demonstrated good reliability, whilst active search for social integration showed acceptable reliability. All other subscales fell short of the acceptable threshold of >0.6. Moderate correlations between 3 items on the willingness to accept help sub-scale were evident, however the remaining items in this scale showed weak correlations. After removing these items, Cronbach’s alpha increased to .71. Therefore, willingness to accept help was retained with the omission of questions 9 and 45. Distance and self-promotion, trivialisation wishful thinking and defence, and finding inner stability were discarded from analysis.

**Table 2.** Reliability scores for each subscale of the Essen Coping Questionnaire.

| *Coping style* | *Cronbach's alpha ($\alpha$)* |
| --- | --- |
| Action, problem orientated coping | .74 |
| Distance and self-promotion | .45 |
| Information seeking and exchange of experiences | .72 |
| Trivialisation, wishful thinking and defence | .30 |
| Depressive processing | .71 |
| Willingness to accept help | .58 |
| Active search for social integration | .69 |
| Trust in medical care | .70 |
| Finding inner stability | .53 |

Acceptable $\alpha = >0.6$

#### HRQoL

At T2, the 11-item Endometriosis Health Profile Questionnaire-5 (EHP-5; Jones et al., 2004) was used to measure endometriosis-specific HRQoL. Five of the six items are scored on a 5-point Likert scale (0: ‘Never’; 4: ‘Always’), whilst the remaining 6 items are scored on the same scale with the addition of a ‘Not Relevant’ option. A composite scale of overall HRQoL was formulated and expressed as a percentage, with 0% representing the best possible HRQoL, and 100% representing the worst. The EHP-5 demonstrated good reliability (α = .79) and was used in both surveys.

#### Procedure

Participants were presented with an information sheet detailing the aims and scope of the survey. They were then asked to confirm that they had received a medically confirmed diagnosis of endometriosis. Participants who answered ‘no’ to this question were directed to the end of the survey. Participants were then asked to provide their contact details so that interested participants could be invited to interview. Participants were debriefed following survey completion. One year later, participants were invited to complete a follow-up survey.

#### Statistical Analysis

Following univariate analysis, a hierarchical multiple regression analysis was conducted to determine whether coping influenced HRQoL, over and above demographic and clinical factors. Demographic data was entered first, followed by clinical endometriosis information, then coping.

### Qualitative Interviews

#### Participants

Thirty participants who had expressed an interest in participating in the interviews within the baseline survey attended online semi-structured interviews. A sampling matrix prioritising the recruitment of individuals with a range of ethnic backgrounds, ages, employment status’, household incomes, and educational attainment was used to ensure a diverse sample (see supplementary material).

Pseudonyms were used to preserve the anonymity of participants. Interviewees were aged between 20-55 years (M = 35.6). Participants had experienced symptoms of endometriosis for between 4 and 40 years (M= 14.83 years, SD= 9.18), and had been diagnosed for an average of 5 years (SD= 6.97). Additional demographic and clinical details relating to participants can be found in table 3.

**Table 3.** Interviewee demographic and clinical details (n = 30)

|  | N | % |
| --- | --- | --- |
| **Ethnicity** |  |  |
| White British | 20 | 66.7% |
| Indian | 2 | 6.7% |
| African | 2 | 6.7% |
| Another Mixed Background | 2 | 6.7% |
| Pakistani | 1 | 3.3% |
| Asian and White | 1 | 3.3% |
| Another Ethnic Background | 1 | 3.3% |
| Another White Background | 1 | 3.3% |
| **Educational attainment** |  |  |
| Attended university | 16 | 53.3% |
| School / college | 14 | 46.7% |
| **Employment status** |  |  |
| Employed full-time / self-employed | 14 | 46.7% |
| Disabled or unable to work | 6 | 20% |
| Employed part-time/ casual contract | 6 | 20% |
| Unemployed | 2 | 6.7% |
| Student | 2 | 6.7% |
| **Country of residence** |  |  |
| England | 17 | 56.7% |
| Scotland | 12 | 40% |
| Wales | 1 | 3.3% |
| **Co-morbid condition** |  |  |
| Yes | 18 | 60% |
| No | 12 | 40% |
| **Surgery** |  |  |
| Had surgery | 24 | 80% |
| Not had surgery | 6 | 20% |

#### Materials

A topic guide incorporating open-ended questions and prompts was utilised for the semi-structured interviews (see supplementary materials). This guide was developed for a broader project on endometriosis-related perceptions (Moore et al., 2023). Participants were asked general questions relating to their endometriosis experiences before several prompts were employed to elicit further information relating to their coping styles, perceptions, and social support.

#### Procedure

The sampling matrix was used to identify potential participants, who were contacted via email in batches of 5. An information sheet was sent to potential participants and those interested provided written consent to be interviewed. Semi-structured interviews were conducted by telephone (n = 5) or online (n = 25), lasting an average of 62 minutes. Interviews were audio recorded. Recordings were deleted following transcription. Participants were debriefed following the interview and offered a £20 Amazon gift voucher as compensation for their time.

#### Qualitative Analysis

Interviews were analysed using Braun and Clarke’s (2021) reflexive thematic analysis (RTA). RTA emphasises the researcher’s active role in knowledge production (Braun and Clarke, 2019) whereby codes are understood to represent the researcher’s interpretations of patterns of meaning across the dataset.

A reflexive journal was kept by the lead researcher throughout analysis to note thoughts and feelings relating to each interview. Transcripts were read and notes taken to aid familiarisation with the data, before the data was distilled into several codes based on the aims of the current study using NVivo v.12. Codes were grouped to construct potential themes, which were reviewed based on their relevance to the research aims. Potential themes were discussed amongst all researchers, before three themes were finalised.

### Integration of data

Morgan’s (2019) approach to integrating mixed-methods research was adopted to combine the qualitative and quantitative components. This approach determines the extent of convergence, complementarity, and divergence across the datasets. By considering these aspects during the integration process, Morgan’s (2019) approach allows for a comprehensive understanding of the data and a holistic perspective of the research area.

## Results

### Longitudinal Survey

#### Descriptive statistics

Participants employed several strategies to cope with the impact of endometriosis. The mean scores for each coping subscale are displayed in table 4.

**Table 4.** Coping strategies used by participants.

| *Coping style* | *Mean* | *SD* | *Range of scale* |
| --- | --- | --- | --- |
| Acting, problem-oriented coping | 1.85 | .69 | 0-4 |
| Information seeking, exchange of experience | 2.00 | .82 | 0-4 |
| Depressive processing | 1.97 | .84 | 0-4 |
| Willingness to accept help | 1.53 | .78 | 0-4 |
| Active search for social integration | 1.50 | .69 | 0-4 |
| Trust in medical care | 1.95 | .73 | 0-4 |

As illustrated in table 4, information seeking and exchanging of experiences was the most used coping strategy, followed by depressive processing, trust in medical care, and action, problem-oriented coping. Participants were least likely to actively search for social integration, for example by meeting new acquaintances, going out with friends, and visiting others, which may be reflective of the social functioning detriments associated with endometriosis (Moradi et al., 2014). As outlined in table 4, no single coping strategy was predominantly employed by participants, indicating heterogeneity in the ways in which participants coped with their experiences of endometriosis.

Participants demonstrated relatively poor HRQoL (M = 58.14, SD = 22.57). There was, however, variation in participant scores as signified by the standard deviation, perhaps reflecting the heterogeneous experiences of individuals experiencing endometriosis.

#### Hierarchical multiple regression

Prior to conducting multivariate analysis, univariate analyses were performed to determine the relationships between each potential predictor and outcome variable, and to establish the factors included within the regression model. Table 5 reveals several statistically significant relationships between participant demographics, clinical status, coping, and HRQoL.

**Table 5.**
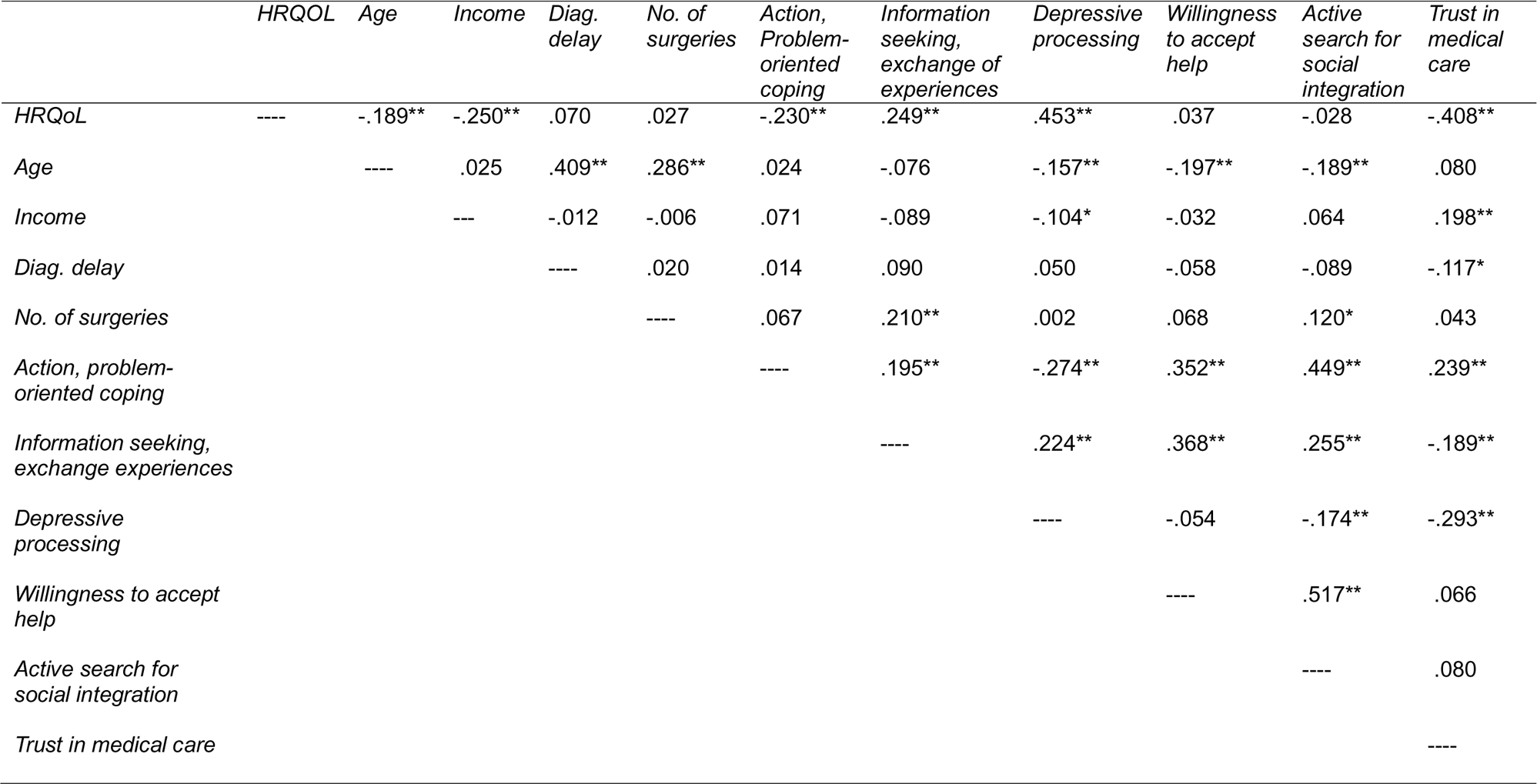
Pearson’s correlation matrix of relationships between predictor and outcome variables.

As demonstrated by table 5, diagnostic delay, surgical history, active search for social integration and willingness to accept help were not related to HRQoL in this sample. Therefore, these variables were dropped from further analysis.

Additionally, a series of t-tests and ANOVAs suggested that educational attainment, employment status, and the presence of a co-morbid condition affected HRQoL. These variables were subsequently included in the final regression model.

Furthermore, as outlined in table 1, most participants identified as from White ethnic backgrounds (94.6%). Therefore, there was too little heterogeneity within the sample to establish whether ethnicity impacted upon HRQoL, and this variable was dropped from further analysis.

A hierarchical regression analysis was next performed to ascertain the impact of coping, demographics and clinical status as measured at T1, on HRQoL, measured at T2 (see table 6).

**Table 6.**
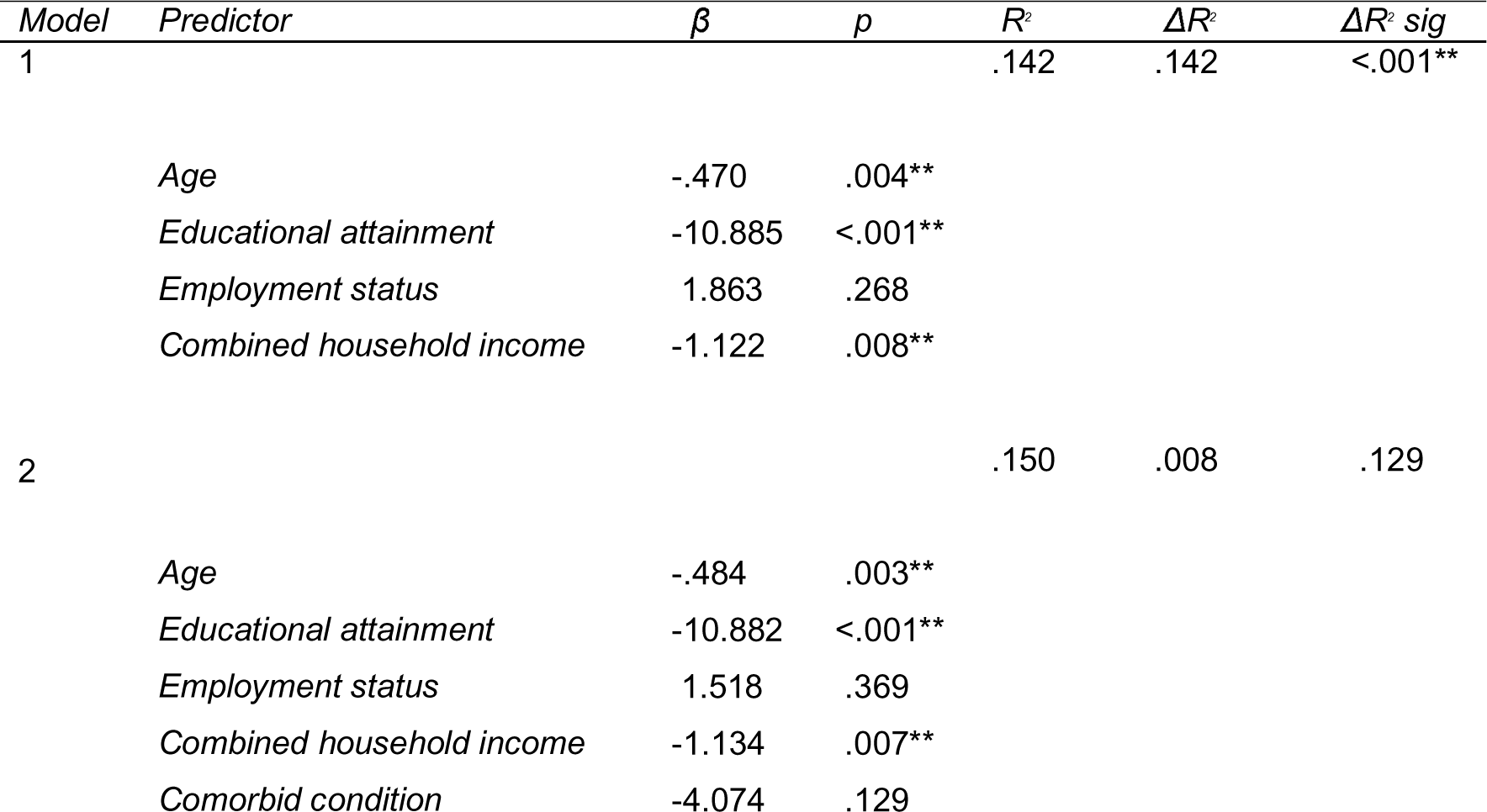
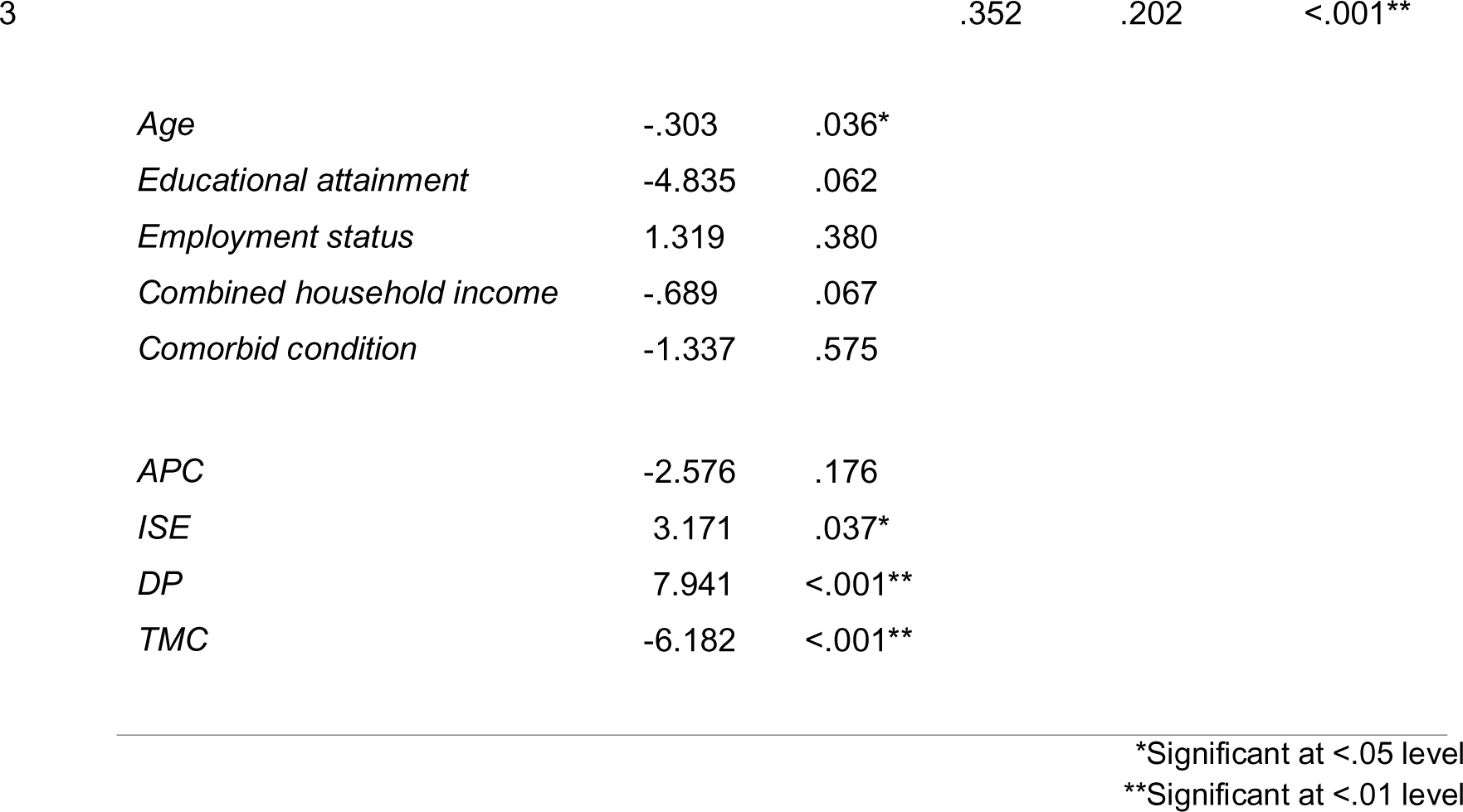
Hierarchical multiple regression results with HRQoL as outcome variable.

The full regression model including demographics, clinical factors, and coping significantly predicted HRQoL, *F*(9, 246) = 14.82, *p*<.001. As illustrated in table 6, the regression model accounts for 35.2% of the variance in HRQoL scores (adjusted R^2^ = .328). As demonstrated by the change in R^2^ in step 3, coping explained 20.2% of the variance in scores over and above demographic and clinical factors.

Considering individual factors, a higher age was linked to more positive HRQoL outcomes. This effect remained even with the addition of coping styles to the regression model. Depressive processing, a coping style characterised by social withdrawal and rumination, and trust in medical care, relating to confidence in medical professionals and treatment, were the strongest individual longitudinal predictors of HRQoL. Heightened depressive processing predicted adverse HRQoL consequences, whilst higher trust in medical care related to more positive HRQoL outcomes. Additionally, information seeking and exchanging of experiences also had a negative effect on HRQoL.

### Qualitative analysis

Three themes relevant to the current paper were constructed through reflexive thematic analysis: i) carrying on and disconnection from the body; ii) balancing boundaries and holistic self-care; and iii) empowered adaptation. Figure 1 provides an overview of each theme before they are considered in greater detail below.

**Figure 1.**
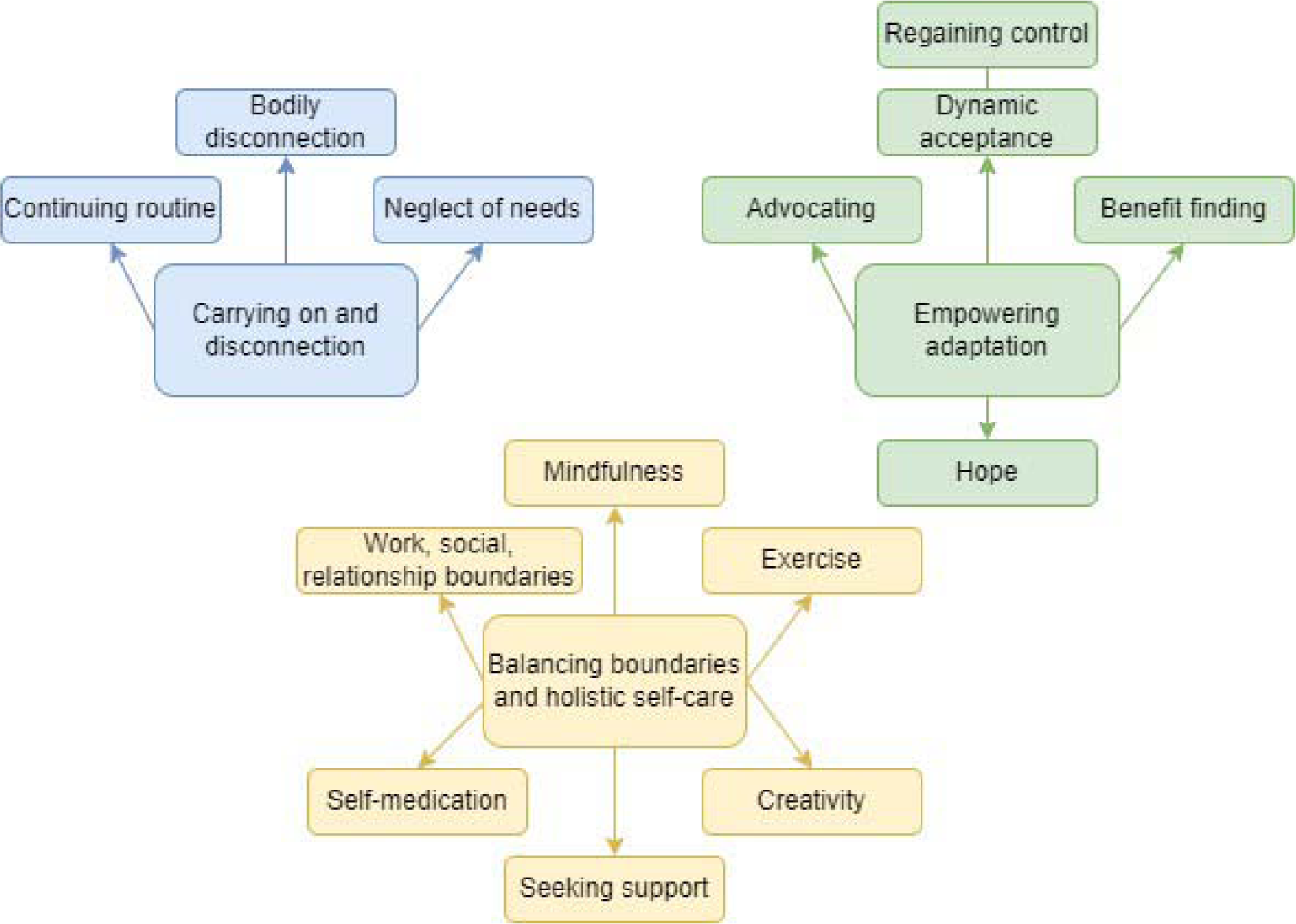
Schematic diagram of themes

#### Theme 1: Carrying on and disconnection from the body

Carrying on and dismissing endometriosis symptoms was a predominant coping mechanism amongst participants. Participants described continuing their usual routine and attempting to ignore their symptoms as far as possible:

> “I know that I just need to, em, get on with it. And I think that’s what always my attitude has been, it’s a case of, this is where I’m at, this is what I need to do. Just get on with it.” [Riley].

Participants often continued with their employment obligations, despite underlying feelings that they needed rest. Such notions were ignored, with work often used as a distraction from these thoughts:

> “I know when I’m at work how little I think about [endometriosis]. So for me yea definitely work is important, it’s an important part of my life.” [Evelyn].

Many participants found comfort in “soldiering on” [Nathalia], maintaining a sense of normalcy and resolving not to let endometriosis dominate their lives. By continuing with their regular routines, participants often experienced a sense of control over their endometriosis and its impact on their lives:

> “…if I’m at work and I look like I shouldn’t be here I’m not here because someone is forcing me to be here, I’m here because I know this is a part of my life and I need to make sure I know how to manage it and deal with it because the option to sit at home and just think about, um, oh my god, you know, the pain, the ovary, the sadness, that’s not an option for me because what kind of life is that?” [Evelyn].

Contrarily, several participants observed that minimising, ignoring, or dismissing their symptoms had unintended consequences, including a sense of disconnection from their bodies. By neglecting to tune in to their bodies’ signals, participants often observed a worsening of symptoms, leading to adverse impacts on their mental and physical health:

> “Rather than be empathetic with myself, I’ve had to push through, I’ve had to turn off all of those emotions of needing rest or needing time or actually, I’m in incredible pain, and just ignore it. And that’s the sad part to me like, actually, you know, that was really a struggle. And I’m so sorry that I had to go through that.” [Reece].

Whilst in the short-term, participants noted that disconnecting from their bodies might provide temporary relief, they also emphasised the significance of re-establishing a connection with their bodies and tending to their needs in the long-term. However, they also recognised the potential drawbacks of this approach. Focussing excessively on endometriosis symptomology, for example, could inadvertently amplify pain sensations and reduce mental wellbeing:

> “I just try and take it day by day, trying not to really focus on ‘oh, it’s really bad’ because then I get really down.” [Iona].

Ultimately, participants stressed the importance of striking a balance between carrying on through their symptoms, and tuning into their bodies to prioritise self-care:

> “I use like coping mechanisms of blocking it out and just forcing myself to get on with things and then I have times where I’m like, no I can’t do this today, it’s not going to work.” [Sarah].

#### Theme 2: Balancing boundaries and holistic self-care

Tuning into their bodies often led participants to set boundaries, particularly with regards to employment (e.g., requesting workplace adjustments), personal relationships (e.g., exploring ways to engage in sexual activity to minimise pain), and social commitments (e.g., rearranging social events). Setting boundaries generally had a positive impact on wellbeing:

> “I’ve learned to say no which is the hardest thing for me. In my work and personal life I’ve noticed it’s benefited me so much more and I’m happier in that sense. So I think that helped my mental health a lot, knowing that it’s okay to say no.” [Jaime].

However, some participants found boundary setting difficult due to the stigmatised and misunderstood nature of endometriosis, which led them to mask their symptoms rather than opening up about their needs:

> “I don’t generally talk about it, um, and that, that makes it harder because then people can’t see, they don’t realise you’re in pain, they don’t know you’re unwell.” [Alex].

Moreover, when setting boundaries, participants often voiced fears of losing relationships and increased isolation:

> “It started impacting my social life whenever I was approaching getting diagnosed I was having to cancel a lot of social events. I ended up losing like a group of people, friends” [Emily].

However, establishing boundaries was crucial for fostering body compassion and self-care, positively impacting participants’ wellbeing. Participants embraced various self-care practices, such as physical activity, mindfulness, and creative outlets:

> “if it’s really, you know, playing with my head I’ll go and sit and I’ll, I’ll meditate and I’ll try and just switch off from it.” [Alex].

> “I draw quite a lot. And see drawing, drawing is therapy. I like, I like to sit and draw when I’m, when I’m feeling down or if I’m sore.” [Billie].

> “I walk a lot. I think being outdoors, um, where it’s quiet and peaceful and beautiful is good for mental health.” [Indra].

Additionally, the self-care techniques employed by participants often involved distraction from painful and distressing endometriosis symptoms:

> “I have to be distracted. I have to be actively distracting myself all the time, um, because of the depression that is made worse by endometriosis” [Jessie].

> And if you can distract yourself from it […] your mind is actually occupied with something else and by the time you know it the painkillers have kicked in and helped the pain go away. [Evelyn].

However, participants’ experiences demonstrate that there is no one-size-fits-all approach to self-care. For some, self-care involved relaxation techniques, such as applying heat through baths or heat pads, while others found comfort in more active and creative outlets, such as exercise or crafts. Importantly, participants noted that different coping styles were employed at different stages of their menstrual cycle. For example, while exercise may not have been feasible during menstruation for some, it served as an important outlet outside of this phase:

> “I do find a little, a little walk, I mean if it’s in the middle of like a really bad flare it’s just not going to happen but if, you know, I’m coming towards the end of my cycle a little walk eases it a little.” [Rowan].

Moreover, forming and nurturing a connection with nature was a particularly prominent self-care strategy amongst participants:

> “I think being close to nature is, it is an escapism, it really is. You’re escaping from everything else in your world that might have you feeling low that day if you’re getting close to nature. It’s amazing, it’s such a good feeling.” [Billie].

Additionally, the importance of tuning in with the body to determine effective coping strategies was emphasised, as described in Theme 1. A minority of participants reported self-medicating with alcohol or cannabis. Whilst alcohol was associated with detrimental effects, such as exacerbating symptoms, cannabis appeared to alleviate pain and symptomology for some individuals:

> “I do smoke cannabis, um, just micro-dose, in very little doses. It helps a lot.” [Jessie].

Ultimately, participants emphasised the importance of developing self-care strategies and self-management strategies that work for the individual.

#### Theme 3: Empowered adaptation

Participants discussed adapting to live a fulfilling life with endometriosis. Seeking social support emerged as an important coping strategy, with participants turning to friends, family, healthcare services, and online groups. Accessing these resources typically reduced isolation and enhanced overall wellbeing:

> “I couldn’t cope without that support group, they’re amazing. They’re an amazing bunch of girls. And you know what makes them more amazing is that they’re going through this too.” [Billie].

Despite the usefulness of support groups in providing support and alleviating feelings of isolation, many participants observed that frequent engagement with these platforms could intensify their focus on pain and the negative aspects of living with endometriosis. Consequently, some participants underscored the importance of accessing these groups in moderation, especially online:

> “Um, [support groups] really helped me, but equally there’s some moments where I have to switch that off, um, because I find that it also has an impact on me so I have to draw a line of how much I read in a day or read someone’s posts” [Jaime].

Nonetheless, openness about their condition frequently led to strengthened friendships and decreased isolation, often giving way to a sense of empowerment:

> “I never used to be very open. But as soon as I started getting really ill I had to start opening up myself. And, emotionally, I’ve gotten a lot closer with friends and family” [Iona].

Further building a sense of empowerment, many participants found purpose in their endometriosis journey by leveraging it to raise awareness and advocate for others. For several participants, finding usefulness in their experiences in this way was important in sustaining their own mental wellbeing:

> “I’m helping other people now, so for me I’m doing this for other people and not for me. So I’m doing something good and it’s sort of kind of making me feel better about myself again.” [Violet].

Additionally, participants emphasised the importance of separating what can be controlled, and what cannot be controlled to support their adaptation to life with endometriosis:

> “I can’t control my body. I can’t control my pain. And I can’t control what’s going on around me. The only thing I can do is control the way I deal with it and the way I live with it.” [Mira].

Adaptation to endometriosis supported some participants to arrive at a point of acceptance in their condition. In this case, acceptance refers to recognising and dealing with the full spectrum of thoughts and feelings associated with endometriosis, rather than trying to avoid or change them. Arriving at a point of acceptance supported the mental wellbeing of participants by reducing apprehension around the future:

> “I think that’s an acceptance thing. Like, if I accept that this might be as good as it gets, um, and, and, know that even if it gets worse, I know better how to cope with it than I ever did before so it’s never going to be as scary and isolating and painful.” [Casey].

However, acceptance in this case was dynamic, in that participants could move in and out of this state:

> “There’s some days where I can accept it and feel okay about [endometriosis], but then there’s days where I’m like, I feel really angry and I feel like devastated and I’m like depressed and all that, like it’s, it’s not linear. It can come and go in all these different stages.” [Emily].

Of those who did reach a point of acceptance, many described a sense of hope that they could live a fulfilling life with the condition:

> “…hope is one of those things that keeps a lot of people going, you know if you have hope you can kind of, you can carry on.” [Becky].

### Integration of qualitative and quantitative results

The qualitative and quantitative results were integrated by considering points of convergence, complementarity, and divergence between the datasets. The results of this process are outlined in table 7 below.

**Table 7.**
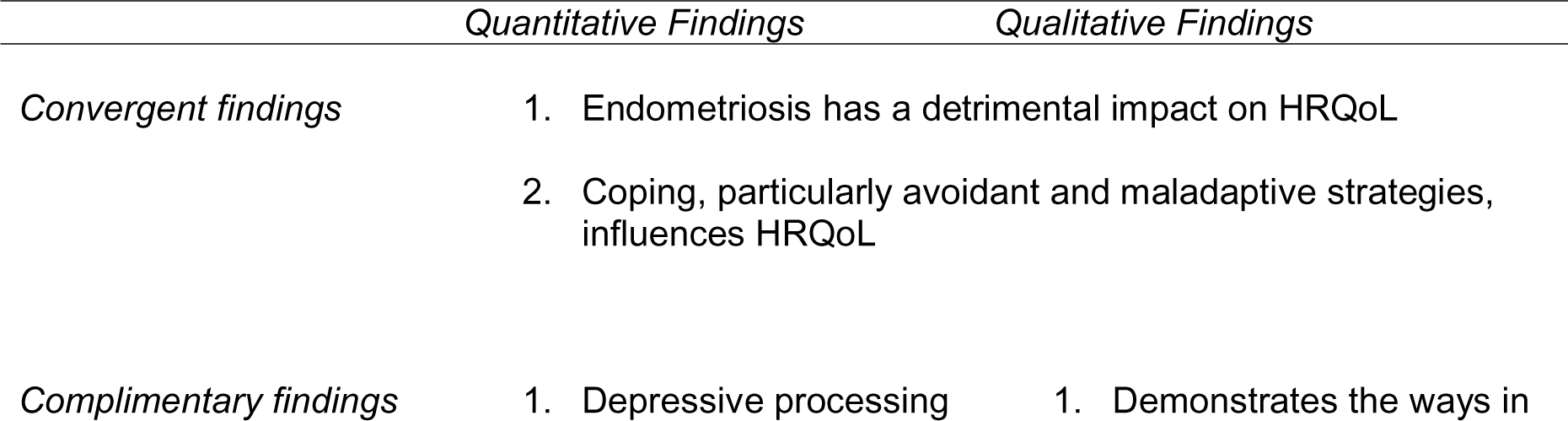
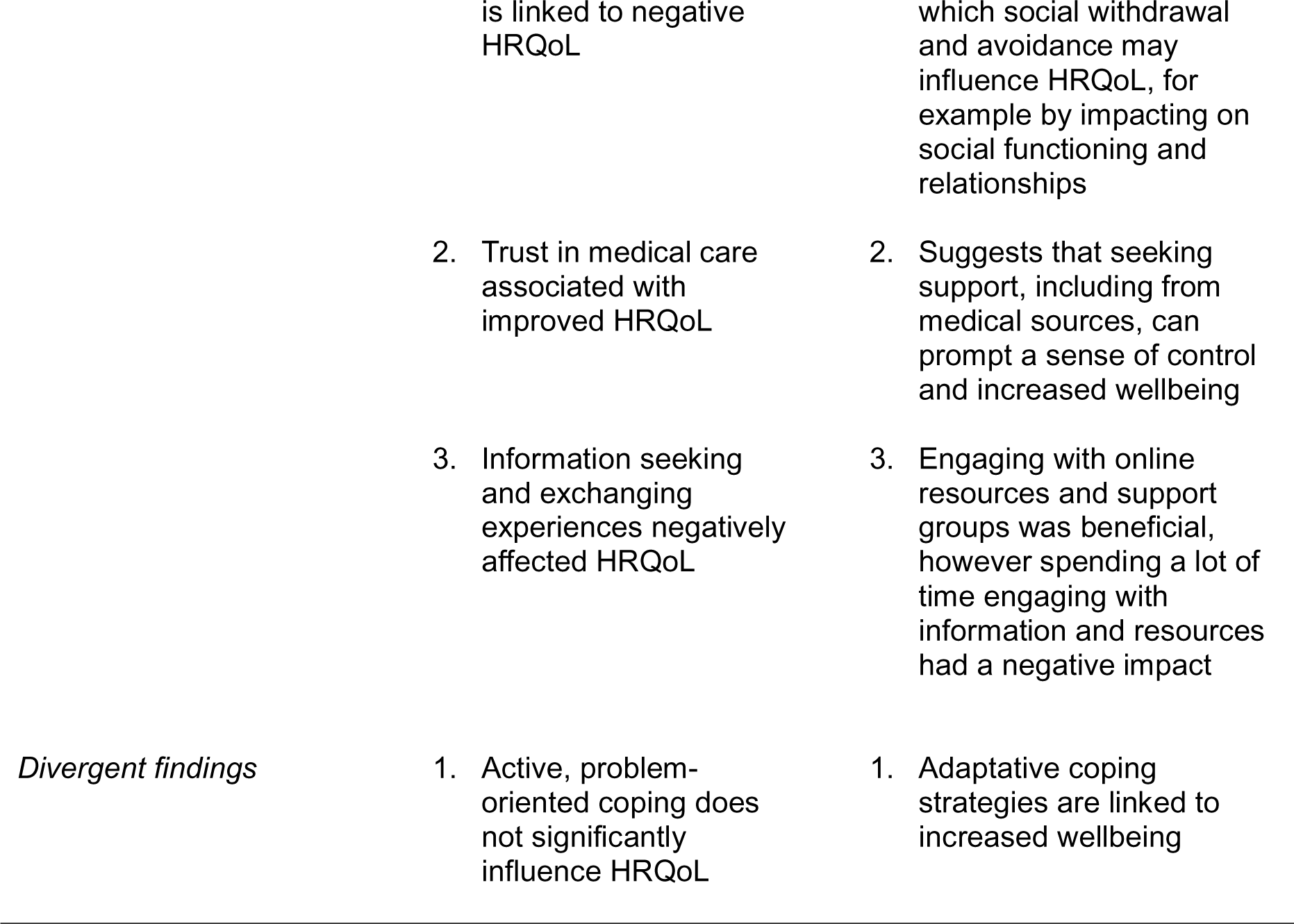
Results of integration of qualitative and quantitative findings.

As outlined in table 7, the integrated findings provide a rich overview of the ways in which coping influences HRQoL in the context of endometriosis. Specifically, the findings converge in relation to the detrimental impact of endometriosis on HRQoL, and the influence of coping in this relationship. The qualitative findings offer complementary insights to aid understanding of some of the quantitative results, by illuminating the mechanisms by which specific coping styles, such as depressive processing, information seeking, and trust in medical care, may impact upon HRQoL. Some divergence between the datasets was observed, however. Specifically, there was no significant influence of problem-orientated coping on HRQoL in the quantitative findings, yet adaption, often incorporating problem-orientated and proactive coping mechanisms, was a predominant theme within qualitative accounts.

## Discussion

The current research is the first to consider the impact of specific coping strategies and adaptation on endometriosis-related HRQoL using a mixed-methods approach. The quantitative and qualitative aspects of the present research largely complemented one another to paint a full picture of the process of adaptation and coping in this population.

More broadly, the findings add to an already sizeable literature base suggesting that endometriosis has a detrimental impact on HRQoL and wellbeing (Wang et al., 2021). The consistent observation that endometriosis is associated with decreased HRQoL and wellbeing suggests a lack of effective psychosocial support for individuals experiencing the condition, and research to aid understanding of the factors underlying reduced HRQoL in endometriosis is therefore imperative to build effective interventions.

Additionally, the findings illustrate the impact of various specific coping strategies on HRQoL and mood. For example, avoidant strategies such as social withdrawal had a marked impact on wellbeing by negatively impacting participants’ relationships, reducing perceptions of control over endometriosis, and increasing isolation. This corroborates the findings of several previous studies, which have linked maladaptive and avoidant coping to decreased QoL in endometriosis (Eriksen et al., 2008; Zarbo et al., 2018). Therefore, interventions to reduce avoidant coping styles, such as cognitive behavioural therapy (CBT) may be useful in this population. At the time of writing, there is limited research on the impact of CBT on coping in endometriosis, however existing research suggests that this kind of therapy may support wellbeing in this population (Evans et al., 2019).

Furthermore, placing trust in medical care had a significant and lasting protective effect on HRQoL. Although trust in medical care has not previously been examined as a coping strategy explicitly in this population, previous research suggests that maintaining a positive relationship with medical professionals can lead to a sense of validation, hope, and positivity in future treatment (Fernley, 2021). Within the current research, participants explained that continuing through endometriosis symptoms and forgoing support may lead to reduced wellbeing, heightened functioning detriments, and disconnection from the body. It follows that confidence in medical treatment and support may lessen these effects, ultimately increasing wellbeing and HRQoL. However, it may be argued that, rather than a coping style, trust in medical care relates to general attitudes towards healthcare. For example, whilst coping typically refers to specific actions or behaviours used to manage stress or adversity, this coping style encompasses beliefs and perceptions about the effectiveness and reliability of healthcare and treatment (Franke & Jagla, 2016). Nonetheless, for some individuals, trust in medical care may indeed function as a coping mechanism, increasing their confidence in treatment effectiveness, encouraging them to seek timely treatment, and instilling a sense of control towards navigating health challenges. Previous research has suggested that relationships between individuals with endometriosis and medical professionals such as GPs and gynaecological specialists are often fraught, marred by widespread misunderstanding within medical settings (Fernley, 2021). Therefore, reparation of this relationship should be prioritised in endometriosis care. Future research may seek to involve both groups to ascertain how this relationship may be repaired, for example by considering the support required by patients, and what may feasibly be offered by medical professionals.

Empowered adaptation emerged as a prominent theme within the qualitative interviews, during which participants stressed the importance of identifying ways to live well endometriosis to support their wellbeing. Adaptation was often intertwined with the identification of self-management techniques and finding usefulness in endometriosis experiences. This observation aligns with the findings of previous research in endometriosis, which has noted a positive impact of adaptation on wellbeing (Yoon et al., 2021). However, problem-focussed coping was not associated with HRQoL within the quantitative element of the current research. At first, this divergence appears counterintuitive, however this observation may be reflective of disparities between the concepts of wellbeing and HRQoL, with the former encompassing broader aspects of psychological and emotional adjustment. Therefore, whilst the adoption of adaptive coping mechanisms may not directly impact HRQoL, it may contribute to overall wellbeing. Future research could investigate the impact of coping strategies on various aspects of wellbeing in endometriosis, for example by considering how different coping strategies influence not only HRQoL but broader dimensions of subjective wellbeing, such as emotional resilience, life satisfaction, and mental health.

Additionally, it is plausible that the quantitative measurement of problem-orientated coping does not sufficiently capture the specific adaptation processes relevant to managing endometriosis. Participants’ responses from the interviews, for example, suggest that adaptation in the context of endometriosis encompasses more than problem-solving, and involves the curation of holistic self-care and coping strategies tailored to living well with the condition. Therefore, the qualitative component of the current research allowed deeper exploration of the multifaceted nature of adaptation, which underlie the divergent results. Indeed, previous research has linked adaptation to chronic illness with disease acceptance, which is a prominent driver of wellbeing in individuals experiencing several chronic conditions (Szcześniak et al., 2020). Therefore, disease acceptance constitutes an important area for future endometriosis research, for example by trialling randomised control trials to ascertain the impact of interventions aimed at supporting disease acceptance, such as acceptance and commitment therapy (ACT).

Aligning with the qualitative findings, previous research has underlined the importance of proactive coping strategies, such as health information seeking, in supporting adaptation to chronic illness (Felton et al., 1984). Information seeking is thought to encourage the adoption of self-care and self-management rituals, leading to increased confidence and even a sense of empowerment (Seçkin et al., 2018). In a study on endometriosis, Sbaffi and King (2020) identified a positive attitude towards information seeking and sharing experiences online. However, this finding runs counter to the findings of the current study, which suggest that information seeking and exchanging experiences has a negative impact on HRQoL and wellbeing. The observed negative impact on HRQoL might be attributed to individuals with more severe endometriosis actively seeking information and support. Additionally, the qualitative findings suggest that seeking out support online was beneficial, but only in moderation. Excessive interaction with online support led to a focus on endometriosis symptomology and worsened pain perception, which may be reflected in the quantitative findings.

Moreover, it is possible that seeking out information for endometriosis may lead to increased distress by highlighting the lack of effective available treatment, the incurable and progressive nature of the condition, and the risk of psychological distress, reflected in lowered HRQoL. At the time of writing, there is no research on the impact of seeking out health literature on wellbeing in the context of endometriosis. To increase understanding of the ways in which information seeking may impact on HRQoL, further research is therefore required. Qualitative research may be most beneficial to ascertain ways of making endometriosis information more accessible and helpful to those with the condition.

### Strengths and limitations

The current research is amongst the first to explore the longitudinal impact of coping on endometriosis-specific HRQoL. By blending quantitative and qualitative methods, it offers a deep insight into the coping processes adopted by individuals with endometriosis, and their impact on HRQoL and wellbeing. Furthermore, by cultivating a holistic account of coping in this population, the insights gained from the current research may be used to create a coping toolkit to support newly diagnosed individuals.

However, the limitations of the current research must also be considered. First, data collection occurred during COVID-19 induced lockdowns, which may have exerted an adverse effect on wellbeing. Lockdowns have been related to limited access to medical treatment (Leonardi et al., 2020) and increased psychological distress in the UK (O’Connor et al., 2021), potentially reflected in participants’ HRQoL scores and interview responses.

Additionally, there was a lack of diversity within the participant sample, particularly concerning ethnicity, necessitating the exclusion of this variable from the quantitative analysis. The underrepresentation of individuals from non-White backgrounds is a prevalent issue in endometriosis research, and serves as the catalyst for a snowball effect whereby the scarcity of data has fuelled misconceptions surrounding the condition (Bougie et al., 2022). Therefore, future research must endeavour to recruit individuals from diverse backgrounds.

A further potential limitation of the study lies in the use of a health-specific measure of coping, which primarily focuses on coping strategies related to health challenges. Whilst this may be valuable for assessing coping styles in the context of endometriosis, it may also overlook more generic coping mechanisms that individuals employ in various aspects of their daily lives, such as managing everyday functioning.

### Conclusion

The current study positions coping as a key driver of HRQoL in the context of endometriosis. Coping had an enduring impact on HRQoL, with the adoption of maladaptive, avoidant strategies exerting a lasting negative impact on HRQoL. Information seeking also exerted a negative impact on HRQoL. Conversely, placing trust in medical care had a protective effect on HRQoL. Participants stressed the importance of adaptation to their condition, by adopting self-care and self-management strategies to support their wellbeing, allowing them often to reach a point of dynamic disease acceptance. Future research should investigate the effectiveness of interventions, such as ACT and CBT, in altering coping responses and improving adaptation to living with endometriosis. Understanding the long-term impact of such interventions could inform the development of targeted approaches to support this population.

## Supporting information

Sampling Matrix

Interview topic guide

## Data availability statement

The data that support the findings of this study are not available due to privacy and ethical restrictions.

## Acknowledgements

This project is funded by a Scottish Graduate School for Social Science Economic and Social Research Council Studentship award (Project Reference: ES/P000681/1). Special thanks to the endometriosis support groups who assisted in the design and recruitment of this research.

