## Supplementary material for "A Mixed-Methods Investigation of Coping, Adaptation and Health-Related Quality of Life in Individuals Experiencing Endometriosis": Sampling Matrix

| **Target demographic** | **Rationale** | **Number to be recruited** |
| --- | --- | --- |
| Non-white ethnicity | Endometriosis has been studied predominantly from the perspective of White individuals due to under-recruitment of people from Non-White backgrounds. Over-recruiting individuals from a diverse range of backgrounds will allow for the experiences of people from different ethnic backgrounds to be heard. | 8-10 |
| Single relationship status | Social support, including intimate relationships, may have a protective effect against some of the QoL-related impact of living with a chronic condition (e.g., Lopez-Martinez, 2008). It is important to consider the experiences of single individuals as well as those in long-term relationships. | 5-8 |
| Not completed University education | Those with University-level degrees tend to be over-recruited in endometriosis research. It is important to consider the views of individuals with a wide range of educational backgrounds. | 5-8 |
| Not in full-time employment | As above, it is important to reflect the views of people in a range of employment situations. In the survey element of the present research, it was noted that most participants were in full-time employment so people in other types of work were under-represented. This is particularly important in this context because people with endometriosis are often unable to work, or work to reduced hours. | 5-8 |
| Over the age of 48 | There is little research around how endometriosis impacts menopausal and peri-menopausal individuals, although recent literature suggests that these individuals may still experience symptoms of endometriosis. It was felt necessary to reflect their views. | 3-5 |
