## Supplementary material for "A Mixed-Methods Investigation of Coping, Adaptation and Health-Related Quality of Life in Individuals Experiencing Endometriosis": Interview topic guide

**Interviewer introduction**

**Any questions?**

**Confirm consent**

**General questions**

**When you think about your experiences of living with endometriosis, how would you say it has affected your life?**

**How do you feel about endometriosis?**

**What thoughts come to mind when you think about your endometriosis and related experiences?**

***Prompts / illness perception category:*** *Relationships? Consequences and impact / support
Working life? Consequences and impact
Home life? Consequences and impact
Self-care and caring for others? Consequences and impact
Emotions toward endometriosis? Emotional representation
Images come to mind? Emotional representation / general IPs
Describe your symptoms/symptom severity? Identity
Understanding of endometriosis? Coherence/identity
Understanding changed since diagnosis? Coherence/identity
Control over endometriosis? Personal control
Control over treatment? Treatment control
How long do you expect symptoms to last? Timeline
Any specific thoughts around the cause of your endometriosis? Cause
Has your diagnosis altered the way you see yourself? Consequences/impact
Strategies to cope? Coping style
Any additional support that could be beneficial? Support*

**Can you tell me how you have found the interview***? Are there things you thought we would have talked about that we haven’t yet? Are there things you would like to share with other people that we haven’t talked about yet?*
